# Predicting 30 Days Hospital Readmission for Heart Failure patients using word embeddings

**DOI:** 10.1101/2025.02.07.25321871

**Authors:** Prabin R. Shakya, Ayush Khaneja, Kavishwar B. Wagholikar

**Affiliations:** Massachusetts General Hospital, Boston, MA USA; Seoul National University, Seoul, South Korea; Havard Medical School, Boston, MA, USA

**Author notes:** Contributorship KBW and PRS conceptualized the study. PRS designed and performed the experiment, prepared the manuscript. KBW reviewed and supervised experiments and manuscript. AK reviewed experiment design and manuscript.

## Abstract

Heart Failure (HF) is a public health concern with a wider impact on quality of life and cost of care. One of the major challenges in HF is the higher rate of unplanned readmissions and sub-optimal performance of models to predict the readmissions. Hence, in this study, we implemented embeddings-based approaches to generate features for improving model performance. Specifically, we compared three embedding approaches including word2vec on terminology codes and CUIs, and BERT on concept descriptions with baseline (one hot-encoding). We found that the embedding approaches significantly improved the performance of the prediction models, and word2vec on the study dataset outperformed pre-trained BERT model.

## Introduction

Heart Failure (HF) is a major and growing public health concern, affecting around 3% of the adult population of developed countries, with increasing prevalence in low and middle-income countries (1,2). It is estimated that 56.2 million people worldwide suffer from HF(2). HF is also associated with high rates of unplanned readmissions, which strain the healthcare system and diminish patient quality of life. The 1-year readmission rate is as high as 53% globally and 59% in the US(3). Given the significant burden imposed by readmissions, healthcare providers strive to identify HF patients at risk for readmission. Several predictive models have been developed for this purpose, including traditional statistical models and machine learning models. However, their predictive performance remains suboptimal in real-world applications, with area under the receiver operating characteristic curve (AUROC) of 0.6 when using electronic health record(EHR) data (4).

Medical codes such as International Classification of Diseases (ICD) codes, procedure codes, and medication identifiers vary in structure and granularity, introducing complexity and high dimensionality into the data. These high-dimensional spaces make it difficult to extract meaningful patterns for prediction. Natural language processing (NLP) and embedding techniques, such as word2vec and BERT, offer a promising approach to address these challenges by transforming discrete medical codes into continuous vector representations. These embeddings capture semantic relationships between medical codes, enabling models to understand the similarities and differences between related diagnoses and procedures (5,6). Several studies have demonstrated that embedding methods may improve the performance of predictive models in healthcare by reducing dimensionality while preserving relevant clinical information (6,7).

To address these challenges, our study, investigates the utility of advanced embedding techniques for improving the prediction of 30-day hospital readmissions in HF patients, leveraging the open data available in MIMIC -IV dataset to develop and evaluate machine learning models. Specifically, we compare two embedding approaches including word2vec on terminology codes and Unified Medical Language System(UMLS) Concept Unique Identifier (CUIs) and BERT on concept descriptors. We hypothesize that leveraging advanced embedding techniques can enhance model performance by capturing complex relationships in high-dimensional medical data, ultimately improving predictions of HF readmissions.

## Methods

### Dataset

For this study, we used Medical Information Mart for Intensive Care (MIMIC)-IV version 2.2, a publicly available dataset released in January 2023. The MIMIC-IV dataset contains comprehensive, de-identified HER data for patients admitted to the Beth Israel Deaconess Medical Center (BIDMC) between 2008 and 2019. The dataset includes 299,712 patients and 431,231 hospital admissions (8).Diagnoses are coded using the ICD, with both 9th and 10th Clinical Modification (CM) revisions. Procedures are recorded using both ICD-9 and ICD-10 Procedure Coding System (PCS) codes. Medications are recorded with the National Drug Code (NDC) along with the Drug name, route, strength, etc. To develop predictive models for HF readmission, we utilized data from the hospital module capturing demographic information, diagnosis, medication and procedure codes.

### Cohort Definition

We identified HF patients using the least restrictive phenotyping (LRP) method, which defines HF-positive cases as any patient with at least one relevant ICD-9 or ICD-10 code for HF (9). We included HF positive cases only for this study. 30-day readmission was defined as any subsequent admission occurring within 30 days of discharge, regardless of the reason for readmission. Patients were excluded from the cohort if they died, or the length of stay was less than one day at the indexed admission. We also excluded data from admissions occurred after the indexed HF admission.

### Data pre-processing

Data preprocessing was structured into three main components: admissions, clinical data extraction, and embedding preparation:

1. **Admissions:** Admissions were categorized as indexed, historic, and future admissions. Indexed admissions were defined as the first hospitalization with a diagnosis of HF. All the admissions prior to indexed admissions were classified historic admission and any subsequent hospitalizations as future admissions. We only developed models for predicting readmission after the index admissions. We included age at admission, length of stay and gender from indexed admissions as features.
2. **Clinical Data extraction:** We extracted diagnoses, procedures, and medications from structured data for selected cohorts. These were split into current if date of record corresponds to the index admission and historic if it precedes the indexed admissions. We applied different encoding or embedding methods to these extracted data. We applied one-hot encoding for baseline. The embeddings approaches applied are described below. The current and historic data are combined with indexed admission features to create the final features sets (fig 1).
3. **Embedding:** We applied three embedding approaches to the feature matrix to capture the sematic representations in the data, to output a new feature matrix.
  A. ***Word2Vec on Terminology Codes:*** ICD codes were concatenated with their version (ICD-9 or ICD-10) to form unique identifiers that account for overlapping codes between the two versions. For medications, we used NDC codes, excluding drugs that lacked NDC identifiers. Each unique code (diagnosis, procedure, or medication) was treated as a word, and the list of codes for each patient encounter was considered as a document. The word2vec model was trained using the skip-gram with negative sampling (SGNS) algorithm. We used the context window size of 5, resulting in 200-dimensional embedding vectors. Separate Word2Vec models were trained for diagnoses, procedures, and medications.
  B. ***Word2Vec on UMLS CUIs:*** To map ICD and procedure codes to Unified Medical Language System (UMLS) CUIs, we implemented a hierarchical string-matching approach. First, codes were matched directly to UMLS using a code-matching algorithm. For unmapped code, we truncated the first three characters and attempted matching again. If multiple CUIs were returned, we selected the most generic code. For Medications, NDC codes were matched and for unmapped codes, we used a string search from the drug name. These mapped CUIs were then treated as ‘word’ and trained in the same manner as recorded medical codes using Word2Vec.
  C. ***BERT on Descriptive Text:*** For each medical code, we utilized its long-form descriptor to generate embeddings using pre-trained BERT models. We employed BioClinicalBERT, which is specialized for clinical texts, to generate context-aware embeddings from the long descriptions provided in the MIMIC-IV datasets. For medication, as long descriptions were not available, we created one by concatenating multiple columns from the prescriptions table. This approach captures semantic meaning and context-specific nuances that are often lost when only the codes are used.

**Fig 1.**
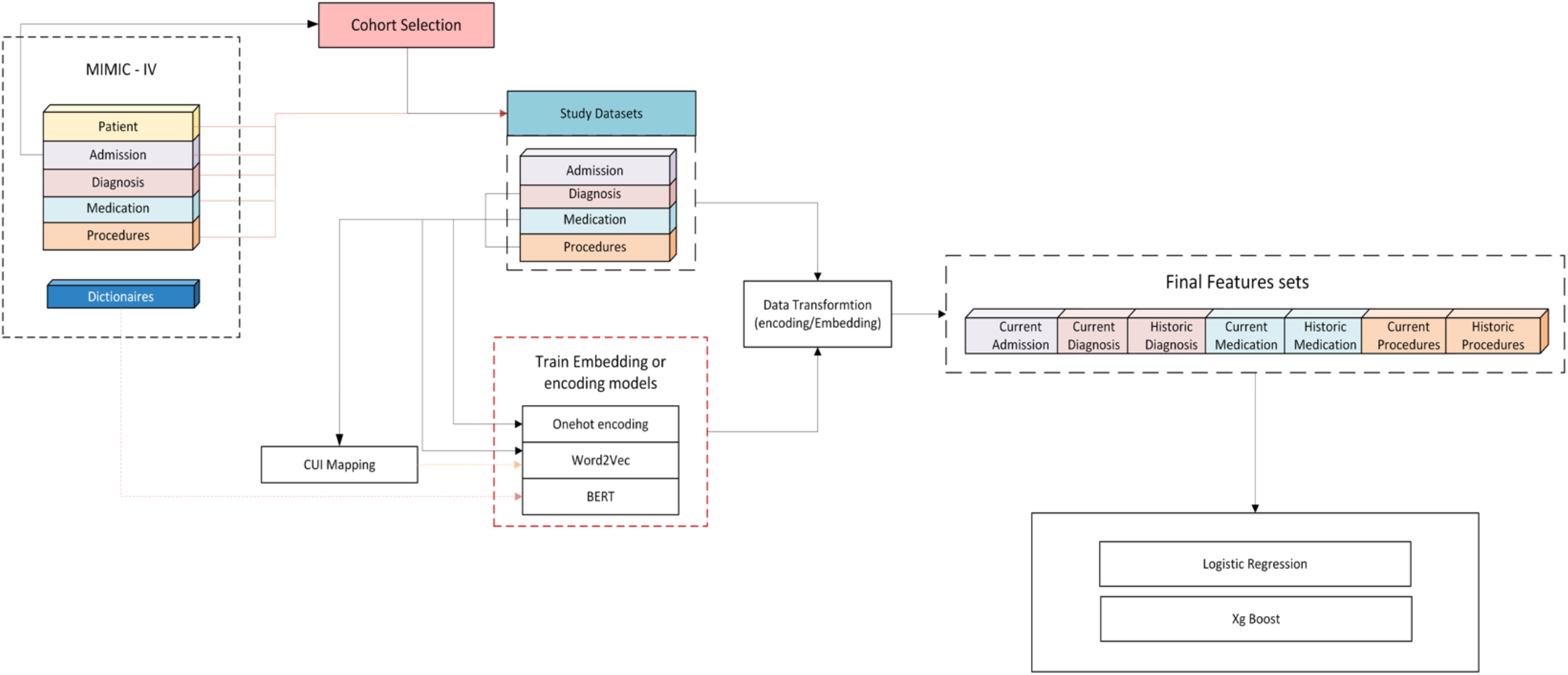
Graphical summary of the methods.

### Model Development

We developed multiple machine-learning models to predict 30-day readmissions. The embeddings generated from each approach were used as input features for these models. We trained and evaluated two machine learning algorithm - logistic regression, and eXtream gradient-boosting (XGBoost). Model performance was assessed using standard metrics such as Area under the receiver operating characteristic curve (AUROC), and F1-score. The source code for model development in this study is available at: https://github.com/dschc/mimicHF_readmission.

### Ethics

The institutional review board at Mass General Brigham determined that the project does not meet the criteria for human subject research, as the dataset used does not contain any identifiable patient information.

## Results

A total of 21,031 patients were included in the study with 3,933 (18.7%) experiencing a 30-day readmission. This cohort was randomly split into training (70%), test (15%) and validation (15%) sets. (fig 2) The average age at indexed admission was 73 years, the mean number of admissions was 2, have mean number of ICD codes used was 25 and the mean length of stay is 8 days. The number of admissions, number of ICD codes, and length of stay are higher among patients with patients with 30-day readmission, and there is no significant difference in age at index admission and the ratio of male vs female. This is true in all splits of datasets as well (table 1). The rate of 30-day readmission was 18% in the Cohort, 19% in the Train dataset, 17% in the test set, and 19% in the validate set.

**Fig 2:**
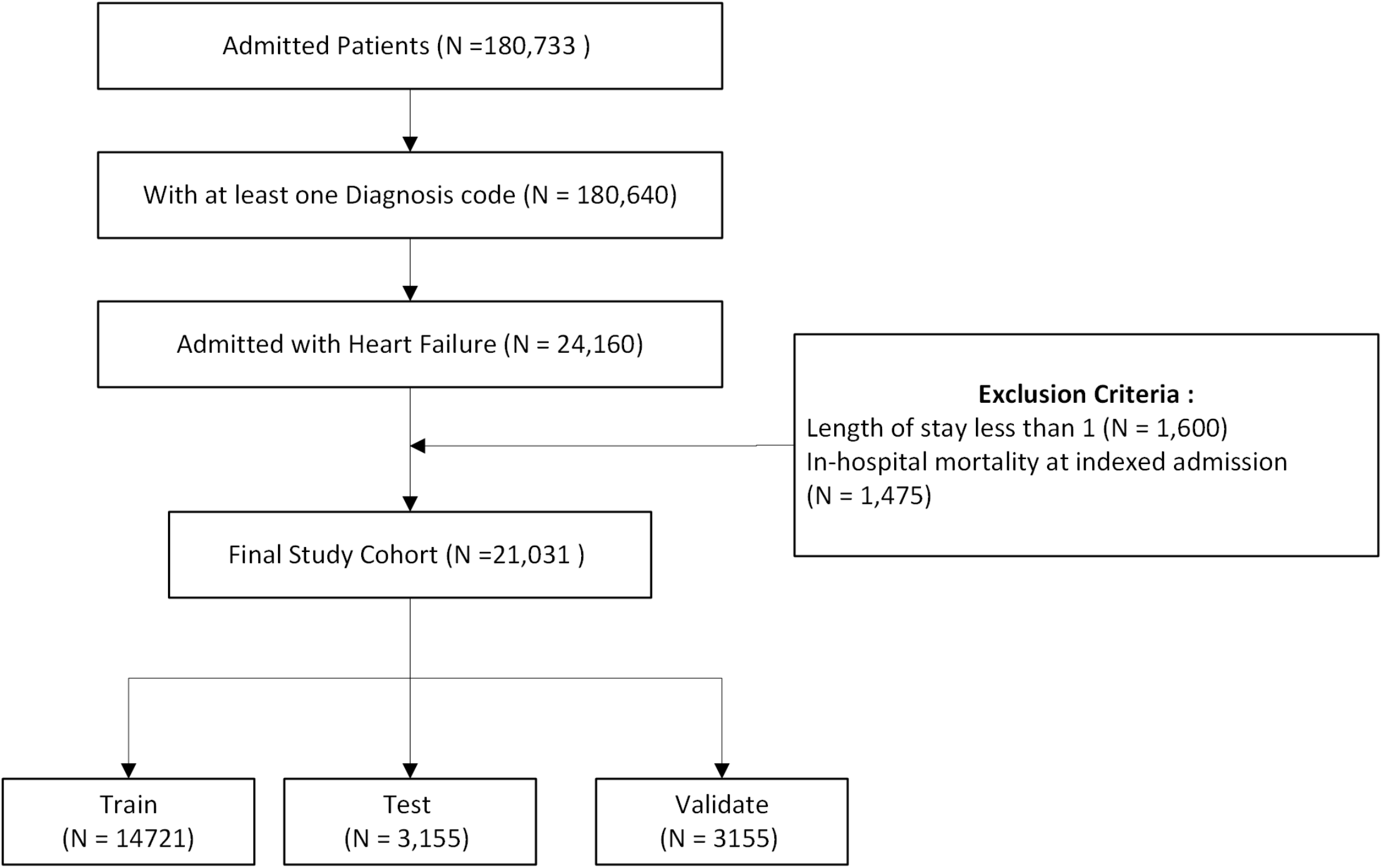
Flow diagram study cohort selection.

**Table 1:** Cohort Characteristics.

|  | Cohort (N = 21031) |  |  |
| --- | --- | --- | --- |
|  | Total | Negative | Positive |
| **Gender** |  |  |  |
| Female | 9971 | 8120 | 1851 |
| Male | 11060 | 8978 | 2082 |
| **Total** | **21031** | **17098** | **3933** |
| # Admissions | 2.07 (2.88) | 1.98 (2.44) | 2.52 (4.28) |
| # ICD Codes | 24.59 (18.12) | 23.83 (16.93) | 27.89 (22.25) |
| Length of Stay | 7.83 (8.57) | 7.53 (8.04) | 9.11 (10.48) |
| Age at index admission | 73.36 (13.55) | 73.39 (13.55) | 73.2 (13.56) |

**Table 2:** Comparison of Model Performance.

| Experiments | Model | AUROC Validation | F1 Validation | AUROC Test | F1 Test |
| --- | --- | --- | --- | --- | --- |
| Baseline | Logistic Regression | 0.54 | 0.26 | 0.53 | 0.25 |
|  | XGBoost | 0.54 | 0.25 | 0.54 | 0.25 |
| Word2vec on Terminology Code | Logistic Regression | 0.64 | 0.37 | 0.64 | 0.35 |
|  | XGBoost | **0.65** | **0.37** | **0.65** | **0.36** |
| Word2vec on CUIs | Logistic Regression | 0.62 | 0.35 | 0.61 | 0.32 |
|  | XGBoost | 0.63 | 0.35 | 0.64 | 0.35 |
| BERT on Descriptive Text | Logistic Regression | 0.59 | 0.32 | 0.59 | 0.30 |
|  | XGBoost | 0.59 | 0.32 | 0.59 | 0.29 |

For each patient, we obtained 52,000 features. We used models with one-hot encoding as the baseline. The baseline AUROC was 0.54 for both the logistic regression and XGBoost models in the validation and test sets, with an F1 score of 0.25. The embedding based approaches performed better compared to the baseline. The AUROC of logistic regression and XGBoost ranged from 0.59 to 0.64 and 0.59 to 0.65 for respectively. Among the embedding methods, BERT on Descriptive Text embedding had the lowest AUROC. The best AUROC was achieved with the word2vec approach applied to terminology codes, using XGBoost (0.65). The F1 scores showed minimal differences, ranging from 0.32 to 0.37 in the validation set and 0.29 to 0.34 in the test set, suggesting that the embedding methods used resulted in only minor performance differences for the prediction models.

## Discussion

The study utilize open EHR data – MIMIC-IV to predict 30-day readmissions for patients admitted with HF. We compared the different embedding methods including embeddings generated utilizing word2vec on terminology codes and CUIs, and ‘BioClinical_BERT’ on descriptor of medical codes. The word2vec based approach treats each medical code as a word and the collection of these codes as a document then uses embedding techniques like word2vec to generate the embeddings(10–13) which captures the semantics of the code. Similarly, second approach which uses pre-trained language models on descriptors of medical codes to generate embeddings demonstrated its capability to capture semantics and relationships, by creating composite vectors using arithmetic functions between vectors(6).

**Fig 2:**
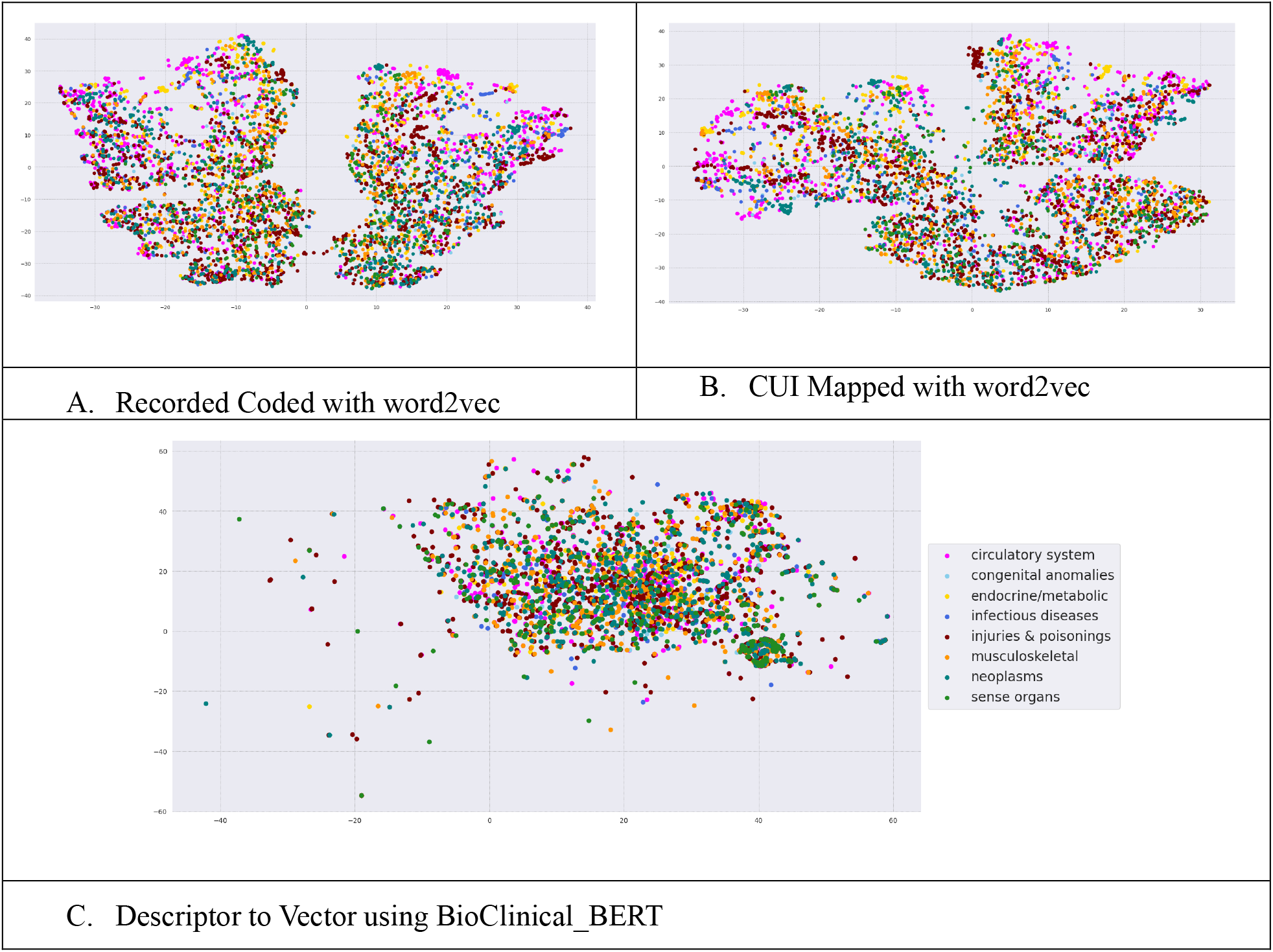
T– SNE plot diagnosis code embeddings with encoded categories.

In our study, we trained the word2vec on training dataset, and adopted bioclinical_Bert trained in medical notes from MIMIC -III(14) which had demonstrated superior performance on NLP tasks. The T-SNE plot of diagnosis codes (ICD-9 and ICD-10) reveals a more coherent representation in the structured EHR-based approach compared to the description-based approach (fig 1). Additionally, model performance was better in word2vec based embeddings, with an AUROC of 0.61 compared to 0.59 and improvement of 0.04 points in the F1 score.

The prediction model performance in the study is in accordance with previous studies(3,7,15,16) which used a data-driven approach of feature generation. The expert-crafted features-based model has higher performance with AUROC greater than 0.8 (17,18). Our study did not explore graph embedding methods, which could serve as promising alternatives. Additionally, we evaluated the performance using logistic regression and XgBoost, which may not fully capture the nuanced differences between embeddings. In the future, we plan to investigate the effectiveness of advanced models, such as deep neural networks, transformers, graphs, and large language models (LLMs).

## Conclusion

The study explored various preprocessing and embedding techniques feature generation-word2vec on terminology code, word2vec in CUIs and BERT on Descriptive Text. The first two approaches utilized the structured EHR data with medical codes, treating the codes as words for generating embeddings. In contrast, the description of the vector employed pretrained BERT ‘bioClinical_BERT’ to represent the medical codes through their text descriptions provided in the dictionary. Word2vec on study dataset outperformed pretrained BERT model in our study. Future work could explore graph-based approaches, LLMs, and neural network-based models for potentially improved outcomes.

## Data Availability

The data is available from physionet.org

https://physionet.org/content/mimiciv/3.1/

## Acknowledgement

We extend our thanks to the developers of open-source tools and frameworks, including word2vec, BioClinical_BERT which greatly facilitated the embedding generation and analysis in our study.

